# Predictors of blood pressure control and antihypertensive drug adherence among hypertensive patients: Hospital-based cross-sectional study

**DOI:** 10.1101/2024.01.11.24301173

**Authors:** Tamrat P. Elias, Asteray T. Minyilshewa, Mengesha A. Tekle, Tsegaye W. Gebreamlak, Binyam L. Adde

## Abstract

**Background:** Hypertension or elevated blood pressure is a serious medical condition that significantly increases the risk of diseases of the heart, brain, kidneys, and other organs. Antihypertensive drug adherence is a key to controlling blood pressure.

**Objective:** To assess factors associated with antihypertensive drug adherence and blood pressure control among hypertensive patients in Selected Public Hospitals under Addis Ababa City Administration.

**Method:** A hospital-based cross-sectional study was conducted among hypertensive patients on follow-up in randomly selected Public Hospitals under the Addis Ababa City Administration from May 1, 2022, to August 31, 2022. The study population was 393 and patients who fulfilled the eligibility criteria were selected by systematic random sampling and the first participant was selected randomly. Data collection was conducted by reviewing the patient’s electronic medical records and by interviewing patients with a structured questionnaire. Data was entered into Epi-Info 7.2.1 and exported to SPSS version 25 software for analysis. Logistic regression analysis was done to see the association between the dependent and independent variables.

**Result:** The rate of antihypertensive drug adherence and blood pressure control was 72.5 % and 23.4% respectively. Participants with uncontrolled blood pressure were found to be 41.7% less adherent than those with controlled blood pressure (AOR= 0.59; 95% CI, 0.36-0.97). Non-adherence to dietary restriction (AOR, 3.31; 95% CI, 1.84–5.96) and chronic kidney disease (AOR=3.85; 95% CI, 1.41-10.52) are associated with good adherence, whereas using single antihypertensive drug (AOR=0.53; 95% CI, 0.30-0.94), and non-adherence to moderate physical exercise (AOR=0.30; 95% CI, 0.20-0.65) were associated with poor adherence to antihypertensive medications. Male sex (AOR=1.95; 95% CI, 1.04–3.28) and blood pressure measurement at home (AOR=0.59; 95% CI,0.36– 0.99) were independent predictors of controlled blood pressure. Drinking alcohol (AOR=1.92; 95% CI, 1.05-3.49) is inversely associated with blood pressure control.

**Conclusion:** Despite good adherence to antihypertensive medications, blood pressure control was low. This signifies the importance of lifestyle measures beyond pharmacologic intervention.

## Introduction

Hypertension or elevated blood pressure is a serious medical condition that can be defined as office systolic blood pressure (SBP) values >=140 mmHg and/or diastolic blood pressure (DBP) values>=90 mmHg, or the level of BP at which the benefits of treatment unequivocally outweigh the risks of treatment [1].

In 2019, an estimated 1.3 billion people Worldwide have high blood pressure, and it is estimated that the number of people with hypertension will increase by 15–20% by 2025, reaching close to 1.5 billion [2,3]. The overall prevalence of hypertension in adults is around 30 - 45% [4]. The estimated pooled prevalence of hypertension is about 30.8% in Africa [5] and 30.0%-31.1% in Sub-Saharan Africa [6]. In Ethiopia, there are different prevalence reports in different studies and across different regions of the country.

Systematic meta-analyses conducted in 2019 using published and unpublished articles showed that the overall pooled prevalence of hypertension in the country was 21.8% [7].

Hypertension is associated with a substantial financial burden. The global financial burden of high BP in 2001 was estimated to be around US$370 billion or about 10% of the world’s overall healthcare expenditure [8].

Elevated BP is also associated with a large global burden of cardiovascular disease (CVD) and premature death. According to data from different studies, an estimated 7.7–10.4 million annual deaths are attributable to elevated blood pressure levels, 88% of which were in low-and middle-income countries (LMICs) [9, 10, 11].

Randomized clinical trials have demonstrated that BP lowering with commonly used regimens, such as diuretics, angiotensin-converting-enzyme (ACE) inhibitors, angiotensin-receptor blockers (ARB), and calcium channel blockers reduces the risk of CVD and all-cause mortality [12, 13].

Despite these effective interventions, hypertension control remains unacceptably low, particularly in LMICs [14]. Recent global estimates suggest that only 45.6% of people with hypertension were aware of their condition, only 36.9% were receiving treatment and only 13.8% had achieved BP control (defined as systolic BP <140 mmHg and diastolic BP <90 mmHg) [14].

In chronic diseases, including hypertension, medication mainly serves as a preventive measure and not to suppress symptoms, maintaining long-term adherence is particularly difficult, and the risk of treatment discontinuation is very high [15].

Various studies have shown that poor medication adherence is a primary reason for poor BP control [16, 17]. Poor adherence is responsible for unnecessary overprescription of drugs, substantial worseningof diseases, increased hospital admission rates, and longer hospital stays all leading to a significant medical burden such as reduced optimal clinical benefit and increased risk of cardiovascular events including acute coronary syndromes, stroke, chronic heart failure as well as mortality [18]. In addition to adverse cardiovascular events, it is also associated with reduced quality of life, disability, reduced work productivity, and greater health care costs [19].

The need of conducting this study is to assess the adherence rate and blood pressure control among hypertensive patients in selected public hospitals under Addis Ababa city administration and identify factors for poor adherence and blood pressure control. So that, knowing the magnitude and reasons for the problem, appropriate interventions can be planned to improve adherence and blood pressure control.

## Methodology

### Study design and setting

A hospital-based cross-sectional study was conducted at Yekatit 12 Hospital Medical College and Menelik II comprehensive specialized hospital medical referral clinics among hypertensive patients on follow-up from May 1, 2022, to August 31, 2022, and the study areas were selected randomly.

Yekatit 12 Hospital Medical College, former Bethsaida Hospital established in 1915, is located in the Arada Sub-city of Addis Ababa. It serves around 230,000 people annually both in the emergency and outpatient departments. Menelik II Comprehensive Specialized Hospital was established in 1884 by the Russian Red Cross Society as a nursing school. Progressively it was named as Menelik II referral hospital and started working under the city government of Addis Ababa Health Bureau.

### Study participants

All hypertensive patients on follow-up at Yekatit 12 Hospital Medical College and Menelik II Comprehensive Specialized Hospital medical referral clinics were used as a source population and patients on antihypertensive treatment were the study population. All patients ≥18 years of age with hypertension and on antihypertensive treatment for at least 6 months were included in the study. The exclusion criteria were refusal to sign informed consent, pregnancy, and known mental illness.

### Data collection tools and procedures

A structured questionnaire and Checklists were developed after a review of relevant works of literature. Several questions that can address the objective of this study were constructed including the independent variables like socio-demographic characteristics (Age, sex, educational status, occupational status, income, marital status, ethnicity, religion, and residence), behavioral characteristics (medication adherence, low salt diet adherence, physical activity status, alcohol status, smoking status), clinical characteristics (duration of hypertension, a family of hypertension, availability of BP cuff at home, BP monitoring at home or elsewhere, comorbidity, number of antihypertensive drugs) and the dependent variables like antihypertensive drug adherence and blood pressure control. The tool was amended after a pre-test was done on 5% of patients having follow-up for hypertension at St. Paul’s Hospital Millennium Medical College. The data was collected by general practitioners working in the chronic medical follow-up clinic of the hospital from May 1, 2022, to November 15, 2022.

### Sample size and statistical analysis

The sample size was determined by using the single population proportion formula. From the study conducted in Dessie Referral Hospital, the overall good adherence rate was 51.9% (P=0.519%) [20]. Since the total population was 4420, a correction formula was used, and a 10% non-response rate was assumed. The final sample size was calculated to be 393. Study participants were selected using systematic random sampling. After the first patient was selected randomly, then every 13th patient who visited the follow-up clinic was selected, until the required sample size was obtained.

After checking the data for consistency, completeness, clarity, and accuracy, it was coded manually, entered into Epi-Info version 7.2.1, and exported to Statistical Package for Social Sciences (SPSS) version 25.0 for analysis. After descriptive statistics, bivariate analysis was done to see the association between the independent and dependent variables. Variables with a P-value of less than 0.25 in the bivariate analysis were further entered into the multivariate logistic regression model for final analysis. Multivariate analysis was done using a backward logistic regression method. P-value less than 0.05 was considered to determine the statistical significance of the association and odds ratio with a 95% confidence interval was used to determine the presence, strength, and direction of association between covariates and the outcome variable.

### Operational definitions

**Hypertension:** is defined as office SBP values ≥140 mmHg and/or DBP values ≥90 mmHg.

**Controlled Blood pressure:** is defined as SBP <130 and DBP < 80.

**Drug adherence:** is defined based on Morisky’s Medication Adherence Scale (MMAS-8) [73]

- Patients were considered as adherent to their medication(s) when Morisky’s medication adherence scale (MMAS-8) score is >6
- Non-adherence was considered when the patient’s MMAS-8 score was ≤6.

### Ethical Consideration

Ethical clearance was obtained from the ethical review committee of Addis Ababa City Administration Health Bureau and the ethical committee of Yekatit 12 Hospital Medical College. Written informed consent was obtained from study participants. Participant identity remains confidential. The answers obtained were documented and analyzed anonymously.

## Results

### Socio-demographic Characteristics

A total of 393 participants were included in the study with a 100% response rate. The majority of the respondents were between 46-60 years (42%) and the mean age of patients was 59.2 (±12.1 SD) years. More than half (57.3%) of study participants were female. About 25% of the respondents completed college/university. Those who live in urban areas constitute the majority of participants (98%) and about 45.8 % of them had a monthly total income greater than 3,000 Ethiopian birr (ETB) per month followed by those who earn 1,000-2,000 ETB (22.4%).

**Table 1:** Socio-demographic characteristics of the patients (n=393).

| **Characteristics** | **Frequency** | **Percentage** |
| --- | --- | --- |
| Sex |  |  |
| Male | 168 | 42.7 |
| Female | 225 | 57.3 |
| Age in years |  |  |
| 18-44 | 48 | 12.2 |
| 45-60 | 165 | 42.0 |
| 61-75 | 146 | 37.2 |
| >76 | 34 | 8.7 |
| Educational level |  |  |
| Unable to read and write | 65 | 16.5 |
| Read and write | 26 | 6.6 |
| Primary School | 67 | 17.0 |
| Secondary School | 68 | 17.3 |
| High School/Preparatory | 68 | 17.3 |
| College/University | 99 | 25.2 |
| Marital Status |  |  |
| Single | 30 | 7.6 |
| Married | 253 | 64.4 |
| Divorced | 25 | 6.4 |
| Widowed | 85 | 21.6 |
| Occupation |  |  |
| Government Employee | 58 | 14.8 |
| Retired | 88 | 22.4 |
| Housewife | 122 | 31.0 |
| Private | 85 | 21.6 |
| No job | 40 | 10.2 |
| Residence |  |  |
| Urban | 385 | 98 |
| Rural | 8 | 2 |
| Monthly income in ETB |  |  |
| <1,000 | 52 | 13.2 |
| 1,000 - 2,000 | 88 | 22.4 |
| 2,001 – 3,000 | 73 | 18.6 |
| >3,000 | 180 | 45.8 |

### Behavioral Characteristics

Of the total 393 participants, 4.6% are active cigarette smokers. The prevalence of alcohol consumption was 22.9%. Most of the respondents ate Vegetables (44%) and Cereal Products (42.7%) over the past 6 months on most days of the week. More than half (59.5%) of respondents use additional salt in their diet. A quarter (75.1%) of respondents had moderate physical activity with a median time of 30 minutes per day.

**Table 2:** Behavioral characteristics of the patients (n=393)

| Behavioral character | Frequency | Percentage |
| --- | --- | --- |
| Cigarette Smoking |  |  |
| Yes | 18 | 4.6 |
| No | 375 | 95.4 |
| Alcohol Consumption |  |  |
| Yes | 90 | 22.9 |
| No | 303 | 77.1 |
| Additional Salt Added on Plate |  |  |
| Yes | 234 | 59.5 |
| No | 159 | 40.5 |
| Vigorous Physical Exercise |  |  |
| Yes | 17 | 4.3 |
| No | 376 | 95.7 |
| Moderate Physical Exercise |  |  |
| Yes | 295 | 75.1 |
| No | 98 | 24.9 |

### Comorbidities

Considering the duration of Hypertension, in almost half (49.6%) of the patients, hypertension was diagnosed within the past 5 years. Diabetes mellitus (49.1%) and heart failure (19.8%) are the two most common Comorbidities. More than one-third of respondents had a family history of hypertension.

**Table 3:**
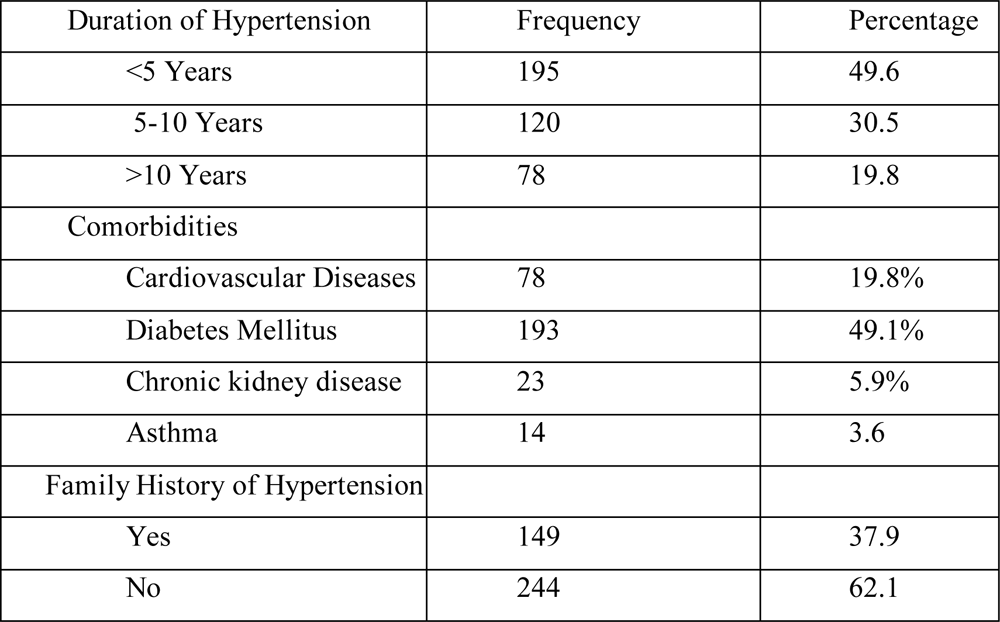
Duration of hypertension and comorbidities of the patients (n=393)

| Duration of Hypertension | Frequency | Percentage |
| --- | --- | --- |
| <5 Years | 195 | 49.6 |
| 5-10 Years | 120 | 30.5 |
| >10 Years | 78 | 19.8 |
| Comorbidities |  |  |
| Cardiovascular Diseases | 78 | 19.8% |
| Diabetes Mellitus | 193 | 49.1% |
| Chronic kidney disease | 23 | 5.9% |
| Asthma | 14 | 3.6 |
| Family History of Hypertension |  |  |
| Yes | 149 | 37.9 |
| No | 244 | 62.1 |

### Antihypertensive Drug Utilization

CCBs (67.2%), ACEIs (58.8%), and Thiazide diuretics (26.5%) are the three commonly utilized antihypertensive drugs. ACEIs and CCBSs are the most common combination treatment accounting for about 33.8%. Monotherapy is utilized in 37.2% of patients, whereas polytherapy is utilized in 62.8% of study participants. Adverse drug reactions were reported in five (1.3%) patients and each of two is due to CCBs and ACEIs and for one patient no specific drug was identified. Most participants (95.4%) bought antihypertensive medication from hospitals and got physician advice about the benefits of antihypertensive drugs.

**Table 4:**
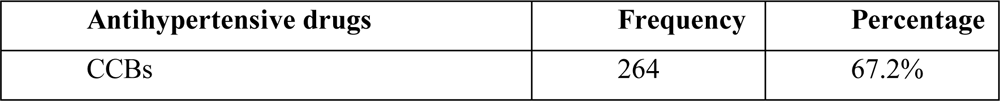
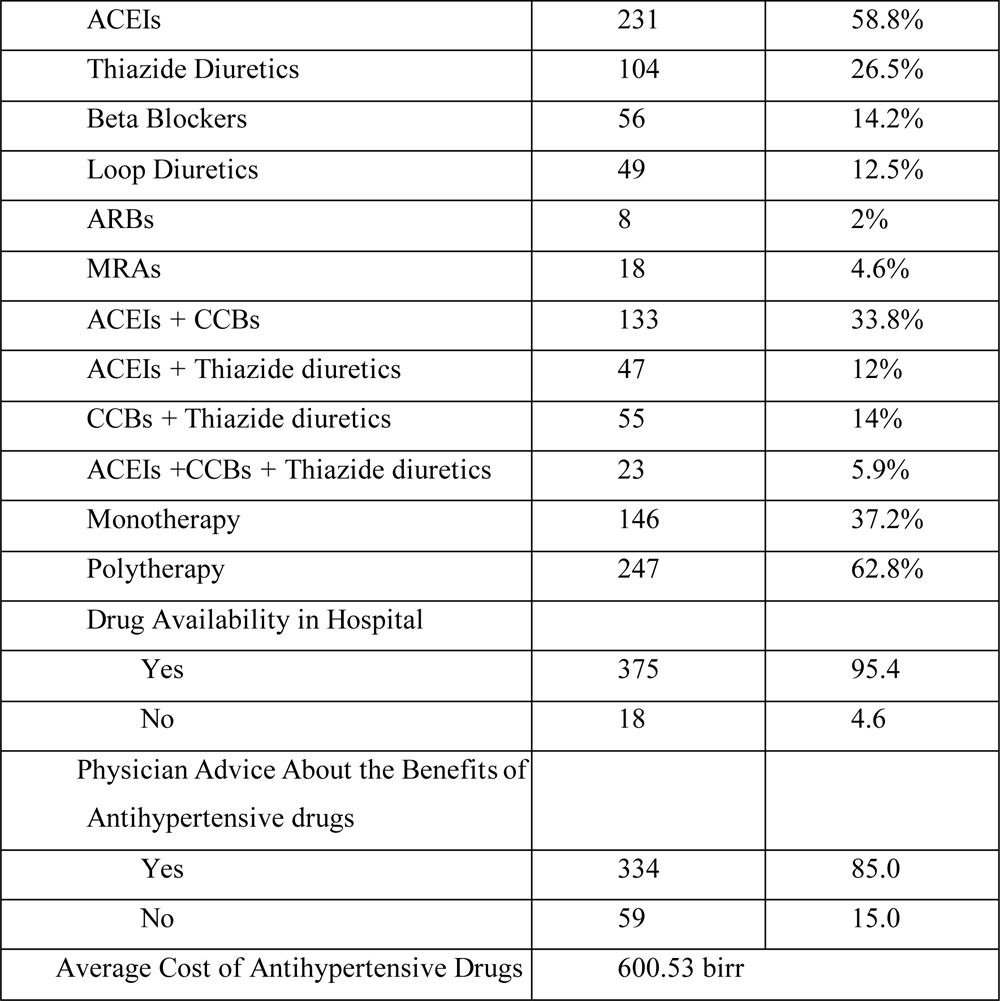
Antihypertensive drug utilization of the patients (n=393)

The respondent’s level of adherence to antihypertensive drugs was assessed using the eight-item Morisky Medication Adherence Scale. It was found that 72.5 % of the respondents were adherent to medication, whereas 27.5 % were not.

More than half (52.4%) of patients measure Blood Pressure at home. The rate of blood pressure control before six months, three months of Study, and at the time of Study was 20.4%, 27.2%, and 23.4% respectively and the rate of recent blood pressure control in Yekatit 12 Hospital Medical College and Menelik II Referral Hospital was 28.8% and 11.5% respectively.

### Factors associated with antihypertensive drug adherence

A multiple logistic regression model was used to assess the association between adherence status and various factors. All statistically significant variables from univariate analysis at a level of 0.25 were included in the multiple logistic regression analysis to control confounding effects. The thirteen factors that were included in the model were sex, age, level of education, monthly income, number of antihypertensive medications, additional salt on a plate, vigorous physical exercise, moderate physical exercise, duration of hypertension, cardiovascular diseases, chronic kidney disease, blood pressure measurement at home, and blood pressure control status.

From the above table, those participants who added additional salt on the plate were 3.3 times more adherent than those who didn’t add additional salt on a plate(odds ratio [OR], 3.31; 95% CI, 1.84– 5.96), those participants who took single antihypertensive medications are 47% less likely to be adherent than those who take two or more antihypertensive medications (odds ratio [OR], 0.53; 95% CI, 0.30-0.94), participants who didn’t do moderate physical exercise were 70% less adherent than those who do moderate physical exercise (odds ratio [OR], 0.3; 95% CI, 0.20-0.65), and participants without chronic kidney disease are 3.8 times more likely to be non-adherent than those with chronic kidney disease (odds ratio [OR], 3.85; 95% CI, 1.41-10.52).

From Bivariate regression analysis, those patients with uncontrolled blood pressure are 41.7% less adherent than those with controlled blood pressure (p=0.041, p <0.05) (odds ratio [OR], 0.59; 95% CI, 0.36–0.97).

**Table 5:**
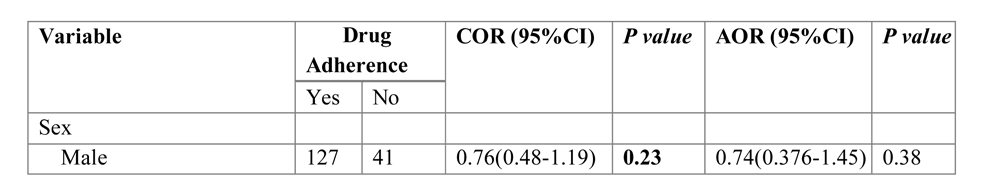
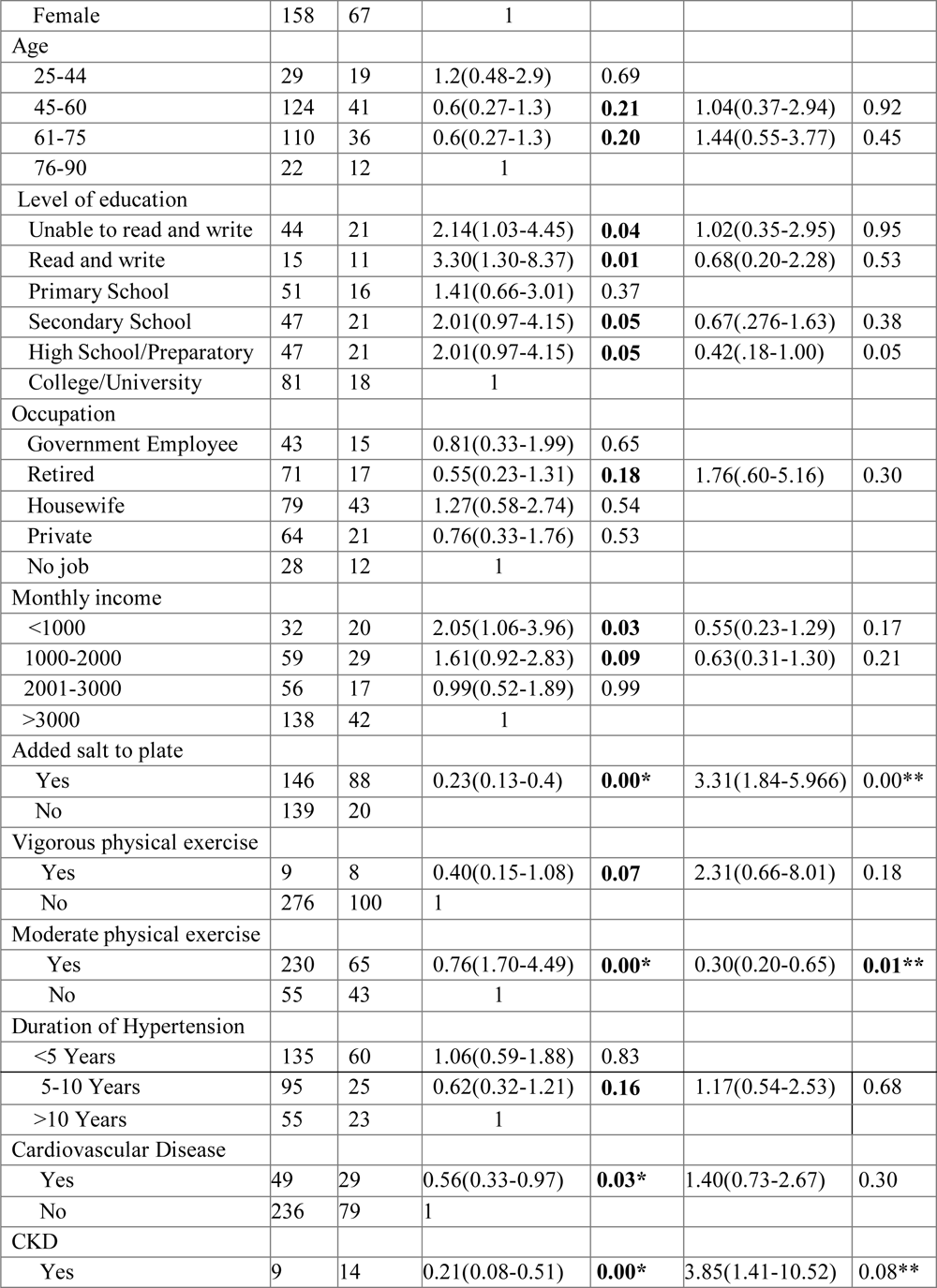
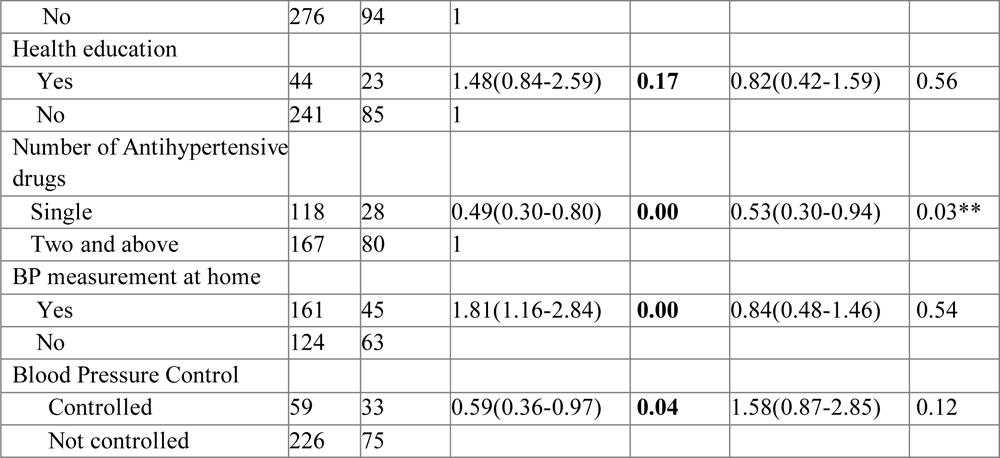
Factors associated with antihypertensive drug adherence.

| Variable | Drug Adherence |  | COR (95%CI) | P value | AOR (95%CI) | P value |
| --- | --- | --- | --- | --- | --- | --- |
|  | Yes | No |  |  |  |  |
| Sex |  |  |  |  |  |  |
| Male | 127 | 41 | 0.76(0.48-1.19) | **0.23** | 0.74(0.376-1.45) | 0.38 |
| Female | 158 | 67 | 1 |  |  |  |
| Age |  |  |  |  |  |  |
| 25-44 | 29 | 19 | 1.2(0.48-2.9) | 0.69 |  |  |
| 45-60 | 124 | 41 | 0.6(0.27-1.3) | **0.21** | 1.04(0.37-2.94) | 0.92 |
| 61-75 | 110 | 36 | 0.6(0.27-1.3) | **0.20** | 1.44(0.55-3.77) | 0.45 |
| 76-90 | 22 | 12 | 1 |  |  |  |
| Level of education |  |  |  |  |  |  |
| Unable to read and write | 44 | 21 | 2.14(1.03-4.45) | **0.04** | 1.02(0.35-2.95) | 0.95 |
| Read and write | 15 | 11 | 3.30(1.30-8.37) | **0.01** | 0.68(0.20-2.28) | 0.53 |
| Primary School | 51 | 16 | 1.41(0.66-3.01) | 0.37 |  |  |
| Secondary School | 47 | 21 | 2.01(0.97-4.15) | **0.05** | 0.67(.276-1.63) | 0.38 |
| High School/Preparatory | 47 | 21 | 2.01(0.97-4.15) | **0.05** | 0.42(.18-1.00) | 0.05 |
| College/University | 81 | 18 | 1 |  |  |  |
| Occupation |  |  |  |  |  |  |
| Government Employee | 43 | 15 | 0.81(0.33-1.99) | 0.65 |  |  |
| Retired | 71 | 17 | 0.55(0.23-1.31) | **0.18** | 1.76(.60-5.16) | 0.30 |
| Housewife | 79 | 43 | 1.27(0.58-2.74) | 0.54 |  |  |
| Private | 64 | 21 | 0.76(0.33-1.76) | 0.53 |  |  |
| No job | 28 | 12 | 1 |  |  |  |
| Monthly income |  |  |  |  |  |  |
| <1000 | 32 | 20 | 2.05(1.06-3.96) | **0.03** | 0.55(0.23-1.29) | 0.17 |
| 1000-2000 | 59 | 29 | 1.61(0.92-2.83) | **0.09** | 0.63(0.31-1.30) | 0.21 |
| 2001-3000 | 56 | 17 | 0.99(0.52-1.89) | 0.99 |  |  |
| >3000 | 138 | 42 | 1 |  |  |  |
| Added salt to plate |  |  |  |  |  |  |
| Yes | 146 | 88 | 0.23(0.13-0.4) | **0.00\*** | 3.31(1.84-5.966) | 0.00** |
| No | 139 | 20 |  |  |  |  |
| Vigorous physical exercise |  |  |  |  |  |  |
| Yes | 9 | 8 | 0.40(0.15-1.08) | **0.07** | 2.31(0.66-8.01) | 0.18 |
| No | 276 | 100 | 1 |  |  |  |
| Moderate physical exercise |  |  |  |  |  |  |
| Yes | 230 | 65 | 0.76(1.70-4.49) | **0.00\*** | 0.30(0.20-0.65) | **0.01\*\*** |
| No | 55 | 43 | 1 |  |  |  |
| Duration of Hypertension |  |  |  |  |  |  |
| <5 Years | 135 | 60 | 1.06(0.59-1.88) | 0.83 |  |  |
| 5-10 Years | 95 | 25 | 0.62(0.32-1.21) | **0.16** | 1.17(0.54-2.53) | 0.68 |
| >10 Years | 55 | 23 | 1 |  |  |  |
| Cardiovascular Disease |  |  |  |  |  |  |
| Yes | 49 | 29 | 0.56(0.33-0.97) | **0.03\*** | 1.40(0.73-2.67) | 0.30 |
| No | 236 | 79 | 1 |  |  |  |
| CKD |  |  |  |  |  |  |
| Yes | 9 | 14 | 0.21(0.08-0.51) | **0.00\*** | 3.85(1.41-10.52) | 0.08** |
| No | 276 | 94 | 1 |  |  |  |
| Health education |  |  |  |  |  |  |
| Yes | 44 | 23 | 1.48(0.84-2.59) | **0.17** | 0.82(0.42-1.59) | 0.56 |
| No | 241 | 85 | 1 |  |  |  |
| Number of Antihypertensive drugs |  |  |  |  |  |  |
| Single | 118 | 28 | 0.49(0.30-0.80) | **0.00** | 0.53(0.30-0.94) | 0.03** |
| Two and above | 167 | 80 | 1 |  |  |  |
| BP measurement at home |  |  |  |  |  |  |
| Yes | 161 | 45 | 1.81(1.16-2.84) | **0.00** | 0.84(0.48-1.46) | 0.54 |
| No | 124 | 63 |  |  |  |  |
| Blood Pressure Control |  |  |  |  |  |  |
| Controlled | 59 | 33 | 0.59(0.36-0.97) | **0.04** | 1.58(0.87-2.85) | 0.12 |
| Not controlled | 226 | 75 |  |  |  |  |

### Factors associated with blood pressure control

The association between blood pressure control with patient-related, medication-related, and Institution-related factors was assessed using bivariate and multivariate logistic regression. Multiple logistic regression models are used to assess the adjusted association of predictors with adherence status. All statistically significant variables from bivariate analysis at a level of 0.25 were included in the multiple logistic regression analysis to control confounding effects. The ten factors that were included in the model were sex, cigarette smoking, alcohol drinking, diet, type of oil used, duration of hypertension, cardiovascular diseases, health education about hypertension and its treatment, drug adherence, and blood pressure measurement at home. Drinking alcohol and male sex and blood pressure measurement at home are independent predictors of controlled blood pressure.

As shown in Table 6, male participants had 1.95 times more controlled blood pressure than female participants (odds ratio [AOR], 1.95; 95% CI, 1.04–3.28), those participants who didn’t drink alcohol had 1.92 times more controlled blood pressure than who drink alcohol (odds ratio [AOR], 1.92; 95% CI, 1.05-3.49) and those participants who didn’t measure blood pressure at home had 41% less controlled blood pressure than those who measure blood pressure at home (odds ratio [AOR], 0.59; 95% CI, 0.36–0.99).

**Table 6:** Factors associated with blood pressure control.

| Variable | BP control |  | COR (95% CI) | P- value | AOR (95% CI) | P-value |
| --- | --- | --- | --- | --- | --- | --- |
|  | Yes | No |  |  |  |  |
| Sex |  |  |  |  |  |  |
| Male | 27 | 141 | 0.3(0.28-0.71) | **0.004\*** | 1.854(1.04-3.28) | **0.035\*\*** |
| Female | 65 | 160 | 1 |  |  |  |
| Cigarette smoking | . |  | . |  |  |  |
| Yes | 91 | 284 | 6.66(0.75-58.8) | **0.08\*** | 0.15(0.01-1.28) | 0.08 |
| No | 1 | 17 | 1 |  |  |  |
| Alcohol Drinking |  |  |  |  |  |  |
| Yes | 63 | 240 | 0.44(0.23-0.83) | **0.01\*** | 1.92(1.05-3.49) | **0.031\*\*** |
| No | 29 | 61 | 1 |  |  |  |
| Diet |  |  |  |  |  |  |
| Meat | 12 | 40 | 1.16(0.47-2.90) | 0.73 |  |  |
| Vegetables | 48 | 125 | 1.75(0.96-3.21) | **0.06\*** | 0.69(0.40-1.21) | 0.20 |
| Cereal Products | 32 | 136 | 1 |  |  |  |
| Type of oil |  |  |  |  |  |  |
| Vegetable Oil | 87 | 276 | 3.22(0.55-18.67) | **0.19\*** | 0.41(0.08-1.94) | 0.26 |
| Butter | 1 | 4 | 2.74(0.14-50.73) | 0.49 |  |  |
| Sesame/Nug Oil | 2 | 5 | 2.30(0.17-30.57) | 0.52 |  |  |
| Others | 2 | 16 | 1 |  |  |  |
| Duration of Hypertension |  |  |  |  |  |  |
| <5 Years | 49 | 146 | 0.66(0.32-1.37) | 0.27 |  |  |
| 5-10 Years | 21 | 99 | 0.49(0.22-1.09) | **0.08\*** | 1.90(0.91-3.96) | 0.08 |
| >10 Years | 22 | 56 | 1 |  |  |  |
| Cardiovascular Disease |  |  |  |  |  |  |
| Yes | 68 | 247 | 0.52(0.26-1.00) | **0.05\*** | 1.77(0.96-3.24) | 0.06 |
| No | 24 | 54 |  |  |  |  |
| Health education |  |  |  |  |  |  |
| Yes | 21 | 46 | 1.68(0.87-3.25) | **0.12\*** | 0.67(0.36-1.25) | 0.21 |
| No | 71 | 255 | 1 |  |  |  |
| Drug adherence |  |  | . |  |  |  |
| Yes | 59 | 226 | 0.56(0.30-1.03) | **0.06\*** | 1.4(0.48-1.46) | 0.21 |
| No | 33 | 75 | 1 |  |  |  |
| BP measurement at home |  |  |  |  |  |  |
| Yes | 56 | 131 | 1.86(1.05-3.30) | **0.03\*** | 0.59(0.36-0.99) | **0.04\*\*** |
| No | 36 | 170 | 1 |  |  |  |

## Discussion

The study showed that the majority of study participants were adherent to antihypertensive medications. But, despite this good adherence, less than one-third of the participants had controlled blood pressure. The addition of salt on the plate, Moderate physical exertion, chronic kidney disease, and the number of antihypertensive drugs were strongly associated with adherence whereas male sex, alcohol drinking, and blood pressure measurement at home were significantly associated with good blood pressure control. Antihypertensive drug adherence was significantly associated with controlled blood pressure.

In this study, the rate of antihypertensive drug adherence was 72.5%, which is higher than other studies conducted in Dessie Referral Hospital (51.9%) [20], Jimma University Medical Center (61.8%) [21], Southwest Ethiopia (60.5%) [22], Nedjo General Hospital (31.4%) [23], and the national medication adherence rate among hypertensive patients (65.1%) [24]. However, the rate of medication adherence was comparable with a study conducted in Sudan, which is 70.5% [25] and lower compared with a study from Tanzania, where the adherence rate was 76.9%, and a study in Japan reported an adherence rate of 91.5% [26].

The variation from other countries could be due to socioeconomic factors and population characteristics and from previous studies in our country could be that most of the studies were done in rural parts where most of the residents didn’t attend formal education. This study is conducted in an urban setup where the society has a better awareness of drug adherence. The other reason could be the difference in the assessment tools.

The rate of blood pressure control was 23.4%, which is lower than the studies conducted in Southwest Ethiopia (50.3%) [22], Jimma University Medical Center (42.8%) [27], University of Gondar Referral Hospital (50.4%) [28], Dessie city (55.8%) [29], Eastern Sudan (45.3%) [25], and Cameroon (36.8%) [30]. However, this study has Comparable results with the US study (24%) [31] which used a similar operational definition of controlled hypertension.

The variation might be due to the difference in the operational definition of controlled blood pressure in previous studies, which was SBP less than 140 and DBP less than 90 as compared to the operational definition in this study, which was SBP less than 130, and DBP less than 80.

A statistically significant association was not found between adherence to antihypertensive medication with patient-related factors including age, sex, level of education, marital status, residence, income, and occupation which is similar to the study conducted at Jimma University Medical Center [27], Eastern Sudan [25], and Cameroon [30].

A statistically significant association was found between adherence to antihypertensive medication and adding additional salt to the plate. Those participants who added additional salt to the plate were more adherent (P<0.01). In this study, the rate of dietary salt restriction was 40% which is higher than reported from two meta-analysis and systematic review studies which was 34% [32] and 37.3% [33]. However, no data report showed an association with medication adherence.

Participants who take single antihypertensive medications were found to be less adherent than those who take two or more antihypertensive medications. Similar findings were reported in Nigeria in which patients taking three or more antihypertensive medications are more adherent with a rate of 73.1% [34] and a study from Lebanon demonstrated taking two or more drugs was a predictor of high adherence (P = 0.02) [35]. Another study from Pakistan also reported that patients on monotherapy had a mean adherence of 79% compared to 90% for those on three drugs or more antihypertensive medications [36]. However, a study conducted at Jimma University Medical Center showed that two or more antihypertensive medication use was associated with poor adherence [21] and a cross-sectional Study conducted in Saudi Arabia showed being on multiple medications was associated with medication non-adherence (P< 0.01) [37].

Chronic kidney disease was associated with good medication adherence in our study. Opposite results were reported from various studies in which the presence of chronic kidney disease was associated with poor drug adherence and the adherence rate also declined with worsening CKD [38, 39, 40]. Of note, a Brazilian study has shown that drug adherence tends to improve as renal function deteriorates [ 41] and another systematic review and meta-analysis showed a 2.54 times increase in the odds of being adherent among patients who had co-morbidities as compared with those without comorbidities [24].

This could be related to physicians and patients becoming more concerned by the quality of BP control as the disease progresses as well as they perceive the combined danger of multiple comorbidities and make them to be more adherent than those without comorbidities.

In this study, male respondents had a higher rate of blood pressure control than females. However, there was an opposite finding from a study conducted in Kenya in which the male sex is an independent predictor of poor blood pressure control [42], a study from the US also reported that male sex was independently associated with uncontrolled blood pressure [43].

Participants who did not drink alcohol had good blood pressure control in this study. REasons for Geographic and Racial Differences in Stroke (REGARDS) Study also showed that heavy alcohol consumption is associated with poor BP control(P=0.02) [40].

Population-based studies conducted in the UK showed higher alcohol use was associated with poor blood pressure control [44].

Participants who measured blood pressure at home had more controlled blood pressure. Similar findings were reported from a systematic review and meta-analysis by Cappuccio FP, blood pressure was lower in people with hypertension who had home blood pressure monitoring than in those who had standard blood pressure monitoring in the healthcare system [45].

Blood pressure control is found to be associated with adherence. Similar findings were reported from several studies demonstrating the strong association between antihypertensive drug adherence and blood pressure control [24, 46–49].

The limitation of this study was the use of indirect methods to assess adherence which predisposes to recall bias due to the self-reporting used for measuring adherence.

## Conclusion

Anti-hypertensive drug adherence is the most important management component for hypertensive patients to achieve adequate blood pressure control which is important in reducing hypertension-associated adverse cardiovascular morbidity and mortality. It was found that 72.4% were adherent to antihypertensive medications despite this only 23.4% of patients had controlled blood pressure. This signifies the importance of lifestyle modifications beyond pharmacologic interventions.

## Data Availability

All data produced in the present work are contained in the manuscript

## Notes

### Competing Interest Statement

The authors have declared no competing interest.

### Funding Statement

This study did not receive any funding

### Author Declarations

Ethics committee/IRB of Yekatit 12 Hospital Medical College gave ethical approval for this work.

